# Prefrontal EEG Delta-Beta Coupling Marks Depressed State in Adult Siblings

**DOI:** 10.64898/2026.10.01.26364438

**Authors:** Mina Elhamiasl, Lilian Y. Li, Felix A. Maldonado Osorio, Brent I. Rappaport, Stewart A. Shankman, Ivan Alekseichuk

**Affiliations:** Stephen M. Stahl Center for Psychiatric Neuroscience, Department of Psychiatry and Behavioral Sciences, Northwestern University Feinberg School of Medicine, Chicago, IL; Department of Psychology, Weinberg College of Arts and Sciences, Northwestern University, Chicago, IL 60611

**Keywords:** Major Depressive Disorder, Electroencephalography, Phase-Amplitude Coupling

## Abstract

Major depressive disorder (MDD) is marked by disorganization in electrophysiological brain networks, yet it remains unclear whether such disorganization reflects the state-dependent pathological effects or a trait-like vulnerability. We analyzed resting-state electroencephalographic (EEG) delta-beta phase-amplitude coupling (PAC) in two sibling cohorts. Current-MDD families included individuals with current MDD and their currently unaffected siblings (n=36), and remitted-MDD families comprised individuals with remitted MDD and their lifetime unaffected siblings (n=110). Delta-beta_high_ PAC was significantly elevated (p_corr_≤0.05) over the left-medial frontal region in participants with current MDD relative to their unaffected siblings. In the remitted-MDD families, no significant differences were observed. Regrouping all individuals irrespective of family ties into current MDD, remitted MDD, and never-depressed controls confirmed significantly elevated frontal delta-beta_high_ PAC at rest in the current-MDD subgroup compared to other subgroups. Overall, delta-beta_high_ coupling reflects the depressive state rather than a trait-like or familial vulnerability.

## Introduction

Major depressive disorder (MDD) is a highly prevalent psychiatric condition characterized by emotional, cognitive, and behavioral disturbances. Rising rates of MDD have been accompanied by a steadily increasing economic burden, with the global economic impact projected to reach US$12 trillion between 2025 and 2050 (Cao et al., 2026). MDD diagnoses are largely based on clinical interviews (Kim et al., 2022). While valuable, these interviews focus on symptomatic rather than mechanistic features and are prone to bias, underreporting, and limited sensitivity (Huang et al., 2023; Kim et al., 2022). Neurobiological biomarkers may help address these limitations by providing objective measures of underlying brain dysfunction that complement symptom-based assessments and offer insight into the mechanisms contributing to MDD.

One such promising mechanistic biomarker is electroencephalography (EEG)-based characterization of neural dynamics. EEG non-invasively measures brain electrical activity with millisecond temporal resolution, providing a direct index of large-scale neural dynamics across distributed brain networks and a powerful tool for investigating altered brain dynamics in depression. A substantial body of research has investigated frequency-band power abnormalities in MDD, identifying alterations across a range of oscillatory frequencies (delta: Dolsen et al., 2017; Schwartzmann et al., 2023; theta: Gheza et al., 2019; Tsai et al., 2023; alpha: Baskaran et al., 2012; Grin-Yatsenko et al., 2010; and gamma: Tsai et al., 2023). Although studies on EEG oscillations have provided important insights into the neurophysiology of depression, most research has traditionally focused on isolated neural processes captured in single frequency bands. Given the increasing evidence that brain function is supported by coordinated activity across distributed neural networks and multiple temporal scales, techniques capable of assessing inter-scale interactions have risen to prominence (Yakubov et al., 2022). Cross-frequency coupling provides such a framework by examining how oscillations in different frequency bands modulate one another, offering a more integrated perspective on the neural mechanisms underlying depression (Raouf et al., 2024). In particular, phase-amplitude coupling (PAC) is the coupling between the phase of a slow, global oscillation and the amplitude of a fast, local oscillation (Canolty & Knight, 2010). PAC represents the coordinated information transfer between the global brain network and a given brain region that underlies complex cognitive and emotional processes (Buzsáki & Draguhn, 2004; Dong et al., 2025; Knyazev, 2007; Putman et al., 2012; Schutter & van Honk, 2005).

Recent PAC studies in mental health highlight delta-beta PAC in particular as both a highly relevant and readily measurable EEG process. Delta-beta PAC has been proposed as a neural marker of state anxiety and stress regulation (Knyazev et al., 2006; Montijn et al., 2023; Schutter & Knyazev, 2012), and has been interpreted as indexing real-time top-down emotion-regulatory processes (Knyazev, 2011). Despite this theoretical relevance, empirical research investigating delta-beta PAC in depression remains scarce. One investigation, conducted in eight individuals experiencing a depressive episode of bipolar disorder, reported decreased delta-beta coupling in the left frontal cortex following three weeks of lamotrigine treatment (Kesebir et al., 2019).

Another study on adolescents undergoing cognitive-behavioral therapy (CBT) for depression showed mixed results: greater post-treatment frontal delta-beta coupling was associated with lower post-treatment self-reported depressive symptoms, while greater post-treatment central delta-beta PAC was associated with higher post-treatment clinician-rated symptom severity (Venanzi et al., 2024). Overall, there is a need for a more systematic analysis of delta-beta PAC in depression. Furthermore, it remains unknown which cortical regions and frequency-band interactions provide the most sensitive and reliable information about depressive pathology.

A further unresolved question is whether delta-beta PAC alterations represent active, state-related processes or reflect a stable, trait-like vulnerability that predisposes individuals to depressive disorders and/or continues into remission. Resolving this distinction is essential for determining whether PAC functions as a biomarker of treatment response and a target for neuromodulation (if it reflects a state) or as a screening indicator of underlying risk (if it reflects a trait). Although the state-versus trait-related nature of delta-beta PAC has not been directly investigated in depression, evidence from stress and anxiety research suggests that delta-beta coupling is sensitive to changes in mental and physiological state. Elevated frontal delta-beta PAC has been associated with enhanced negative perceptual bias and stress reactivity during stress anticipation (Montijn et al., 2023). Similarly, delta-beta PAC was associated with state nervousness during early anticipation of a stressful situation, although this association was observed only among individuals with low social anxiety (Poppelaars et al., 2018). One study found that painful stimulation increased delta-beta coupling, with greater coupling also associated with behavioral inhibition in anxious individuals (De Pascalis et al., 2020). During performance evaluation, higher state anxiety was also associated with greater delta-beta amplitude-amplitude and phase-amplitude coupling in the orbitofrontal and anterior cingulate cortices (Knyazev, 2011). In another study, however, a greater cortisol response after a social stress task was associated with delta-beta amplitude-amplitude but not phase-amplitude coupling (Wang et al., 2024). Evidence from physiological manipulations further supports the state sensitivity of delta-beta coupling. The correlation between delta and beta power increased following cortisol administration (van Peer et al., 2008), whereas it decreased following testosterone administration, which inhibits stress-induced hypothalamic-pituitary-adrenal axis activation (HPA) (Schutter & van Honk, 2004), and following psychotherapy in individuals with social anxiety (Miskovic & Schmidt, 2009). Together, these findings suggest that delta-beta coupling can reflect transient changes in psychological and physiological state.

Whether delta-beta PAC similarly varies as a function of depressive state, however, remains unknown. Several approaches can be used to disentangle state and trait effects, including experimental manipulation of psychological or physiological states and trait-state-occasion modeling (see Rappaport et al., 2025). Within depression research, two informative and practical approaches are family-based designs and current-versus-remitted cross-sectional designs. Comparing individuals with current depression and those in remission is widely used; however, current-versus-remitted designs struggle to separate state effects from selection effects. Not all current patients will remit, and thus any identified group differences might be due to either the depressive state or a trait-like predisposition to remit. Family-based designs partly circumvent this issue because siblings share about half of their genes and much of their environment. A marker that differs between a currently depressed individual and their non-depressed sibling, yet does not differ between a remitted individual and their never-depressed sibling, is more plausibly tied to the depressive state than to familial vulnerability (and vice versa).

To address the gaps in understanding EEG network coordination in depression and its specificity to depressive state versus trait, we examined resting-state delta-beta PAC using a sibling design comprising two cohorts: (a) current-MDD families, in which one sibling was experiencing a current depressive episode while the other was currently asymptomatic, and (b) remitted-MDD families, in which both siblings were currently asymptomatic, but one had a history of MDD and the other had no lifetime history of depression. We hypothesized that delta-beta PAC would be abnormal in the MDD groups and aimed to arbitrate its state versus trait nature. If it reflects a trait, it could inform screening and patient stratification. If it reflects a state, it could serve as a biomarker for monitoring symptoms and treatment response and for personalizing interventions.

## Methods

### Participants

Participants were drawn from a larger family study examining transdiagnostic mechanisms of psychopathology (see Gorka et al., 2016) for detailed recruitment procedures). In brief, sibling pairs between 18 and 30 years old were recruited through community and clinic advertisements in the Chicagoland area. Individuals with severe internalizing psychopathology and alcohol use were oversampled. All procedures complied with the Declaration of Helsinki and received approval from the Institutional Review Board (STU00209363).

The current analyses included 146 participants (73 sibling pairs) divided into two independent cohorts of families based on MDD status determined by the Structured Clinical Interview for DSM-5 Disorders (First et al., 2015). The current-MDD families consisted of 36 participants (18 sibling pairs), in which one sibling met criteria for current MDD, and the other sibling was not currently depressed (either never depressed or currently remitted). The remitted-MDD families included 110 participants (55 sibling pairs) without current depressive symptoms, in which one sibling had a past history of MDD, while the other sibling had no lifetime history of MDD. Sample size was determined by the available recruited pool of sibling pairs meeting inclusion criteria; a post hoc sensitivity analysis indicated that the current-MDD families provided ≥ 80% power to detect within-pair effects of Cohen’s d ≥ 0.70 at α = 0.05 (two-sided), and the remitted-MDD families provided ≥ 80% power to detect effects of d ≥ 0.38. Participants’ demographic and clinical data are summarized in Table 1.

**Table 1.**
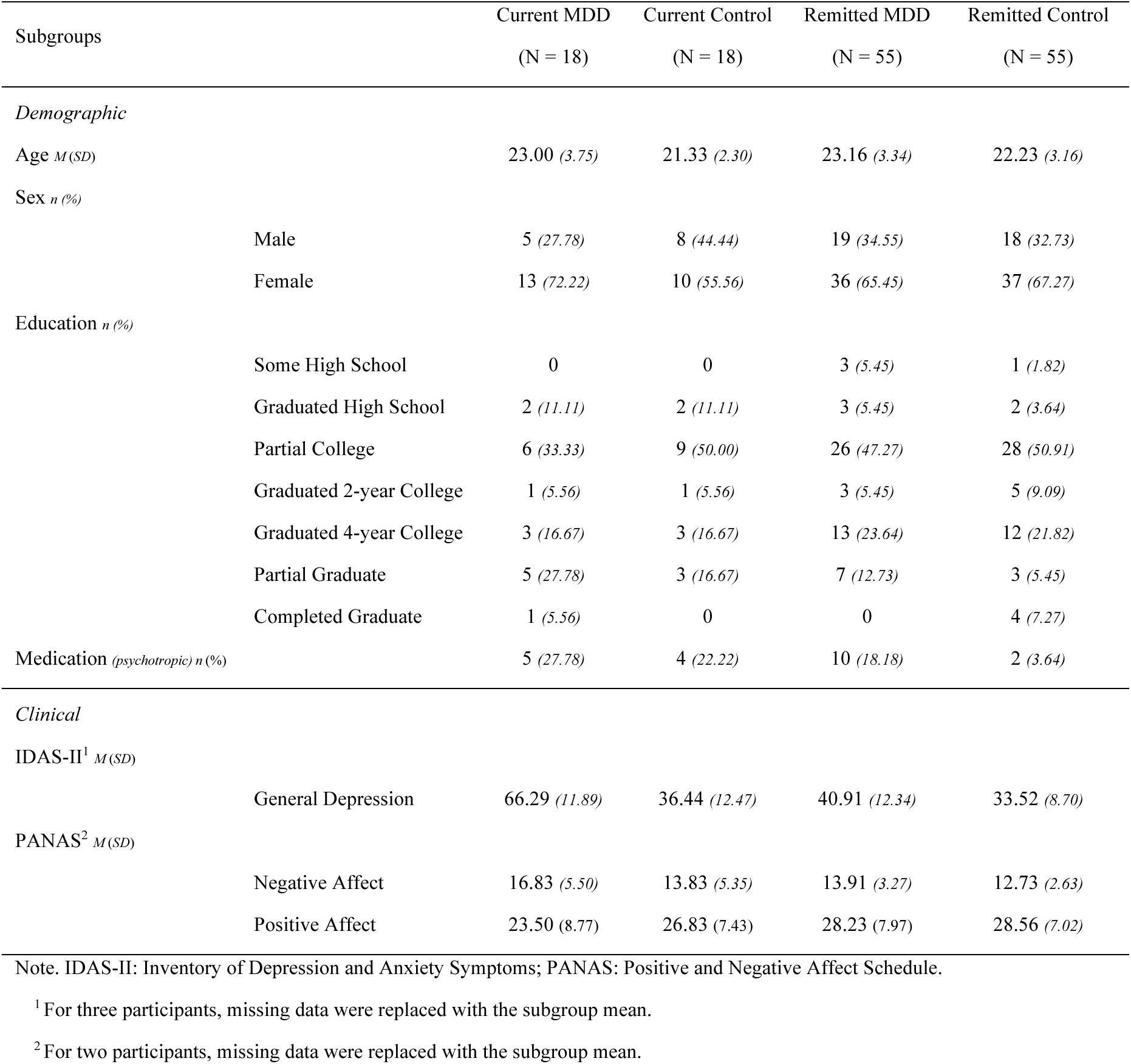
Participant demographic and clinical characteristics.

### Self-Report Measures

#### Positive and Negative Affect Schedule (PANAS)

(Watson et al., 2012). The PANAS was used to assess current affective state. It consists of two 10-item subscales measuring positive affect (PA; e.g., enthusiasm, alertness, and engagement) and negative affect (NA; e.g., distress, fear, and irritability). Participants rated how strongly they experienced each emotion on a 5-point Likert scale ranging from 1 (very slightly or not at all) to 5 (extremely). Scores for each subscale were calculated by summing the corresponding items, with higher scores indicating greater levels of positive or negative affect. The PANAS demonstrated good internal consistency in the present sample, with Cronbach’s α = 0.885 for PA and α = 0.762 for NA.

#### Inventory of Depression and Anxiety Symptoms-II (IDAS-II)

(Watson et al., 1988). The IDAS-II is a multidimensional self-report measure designed to assess symptoms associated with depressive and anxiety disorders. In the present study, the General Depression subscale (20 items) was used to quantify overall depressive symptom severity. Participants rated the extent to which they experienced each symptom using a 5-point Likert scale ranging from 1 (not at all) to 5 (extremely). Scores were summed across items, with higher scores reflecting greater severity of depressive symptoms. In the present sample, the General Depression subscale demonstrated good internal consistency, with a Cronbach’s α of 0.892.

### EEG Data Processing

A three-minute continuous EEG recording was obtained in the eyes-closed condition. Data were acquired using the BioSemi ActiveTwo system (BioSemi, Amsterdam, Netherlands) at a sampling rate of 1024 Hz. The recording montage included 64 Ag/AgCl electrodes positioned according to the standard 10-20 layout. The EEG recordings were referenced online to a common mode sense active electrode positioned between PO3 and POz, along with a ground electrode located between POz and PO4. Electrode offsets were maintained within ±40 mV to ensure optimal signal quality. Online filtering of the EEG data employed a fifth-order low-pass sinc filter with a 3 dB cutoff frequency set at 208 Hz (Li et al., 2023, 2025).

EEG data preprocessing was conducted using MATLAB R2025b (MathWorks, Natick, MA) in combination with the FieldTrip toolbox (Oostenveld et al., 2011). Raw continuous EEG recordings were band-pass filtered at 0.1-55 Hz using fourth-order IIR filters. Bad or noisy channels were visually inspected, identified, and interpolated based on neighboring electrodes. The average number of interpolated channels was 0.49 per recording. Independent Component Analysis (ICA) was applied to the filtered and interpolated data to decompose the EEG signals into independent components, allowing for the manual identification and removal of artifacts such as eye movements, muscle activity, and line noise. The average number of removed components was 0.36 per recording. After ICA correction, the data were re-referenced to the common average. The spatial distributions of delta (2-4 Hz), low-beta (beta_low_ =13-20 Hz), and high-beta (beta_high_ = 20-30 Hz) activity across all electrodes for each subgroup are shown in Figures S1 and S2 in Supplementary Materials.

### Phase-Amplitude Coupling (PAC)

Within-channel PAC was calculated to quantify cross-frequency interactions between low-frequency phase and high-frequency amplitude components of the EEG signal. Given prior findings showing differential associations of beta_low_ and beta_high_ activity with psychological conditions such as MDD, we calculated and examined phase-amplitude coupling separately for delta-beta_low_ and delta-beta_high_ bands (De Filippi et al., 2021a; Hao et al., 2025; Johnson et al., 2026; X. Liu et al., 2024). Preprocessed and re-referenced EEG data were used for these analyses. To obtain the phase and amplitude time series required for PAC, the continuous EEG data were first band-pass filtered separately for the low-frequency phase-providing band (2-4 Hz delta) and the high-frequency amplitude-providing band (13-20 Hz beta_low_ and 20-30 Hz beta_high_) using finite-impulse-response (FIR) filters with a one-pass, zero-phase design to avoid phase distortions. The instantaneous phase and amplitude of the filtered signal were obtained from the Hilbert transformation. Continuous data were segmented into overlapping epochs of four seconds with 75% overlap to enhance the stability of PAC estimates. PAC values were quantified using the modulation index (MI), which measures the extent to which the amplitude of the high-frequency signal (beta) is modulated by the phase of the low-frequency signal (delta) (Canolty et al., 2006). For each channel, MI was computed across all epochs, producing trial-level measures averaged into channel-level MI measures. Complementary metrics, including delta and beta power and delta phase autocorrelation, were also extracted to characterize oscillatory dynamics and MI confounds. To control for spurious correlations in the MI estimates, surrogate distributions were generated by randomly reshuffling the amplitude time series across the trials for 1,000 iterations. Observed MI values were then normalized by z-scoring against the surrogate mean and standard deviation. All analyses were conducted separately for delta-beta_low_ (2-4 Hz phase × 13-20 Hz amplitude) and delta-beta_high_ (2-4 Hz phase × 20-30 Hz amplitude) PAC to assess frequency-specific cross-frequency coupling effects.

Following the whole-head PAC analyses, we examined the comodulogram of phase-amplitude coupling at the center of the significant cluster (the left frontal electrode F3) to better characterize the spectrum of PAC. Preprocessed and re-referenced EEG data were extracted using the FieldTrip toolbox, and the signal was segmented into four-second epochs with 75% overlap. PAC was computed across a grid of phase frequencies (1.25-5 Hz) and amplitude frequencies (13-30 Hz) using the modulation index (MI) as described above. Within-electrode normalized PAC values were calculated for each phase-amplitude pair.

### Statistical Testing

#### Cluster-based statistics of topographic PAC

Within-subgroup Wilcoxon signed-rank tests were used to identify electrodes at which MI values were significantly higher than spurious levels (from the surrogate distributions) in the current MDD, current control, remitted MDD, and remitted control subgroups. For both current-MDD and remitted-MDD families comparisons, Wilcoxon signed-rank tests were also conducted to determine the between-subgroup differences. In the next step, cluster-based permutation analysis was used to investigate significant clusters of electrodes. Cluster-based analysis increases statistical sensitivity by assuming that neural effects occur in spatially contiguous groups across relevant dimensions such as space, time, and/or frequency (Candia-Rivera & Valenza, 2022). From the Wilcoxon signed-rank tests performed at each electrode, we extracted the resulting test-statistic values for each participant and converted their p-values into a binary significance map (1 = significant electrode, 0 = non-significant). These two vectors served as the inputs to the subsequent cluster-based permutation analysis. Clusters were defined as spatially neighboring significant electrodes, requiring a minimum cluster size of three electrodes. For each cluster, we computed a cluster statistic based on cluster mass (sum of the absolute test statistics across all electrodes in the cluster). To evaluate the statistical significance of the observed clusters, we generated a permutation-based null distribution (5,000 permutations). In each permutation, the number of significant electrodes was preserved by randomly shuffling electrode labels and reassigning the same number of significant positions. Because our analysis used the cluster-mass metric, the corresponding test-statistic values were shuffled in the same manner. For each permuted map, we identified all clusters that met the minimum size criterion and recorded the maximum cluster-mass value, yielding a max-statistic null distribution across 5,000 permutations (Phipson & Smyth, 2010). Clusters with p < 0.05 (two-sided, cluster-level corrected) were considered significant.

#### Cluster-based statistics of PAC comodulogram

Following identification of the significant electrode cluster, we analyzed the PAC comodulogram at the center of the significant cluster (F3 electrode) to further characterize the frequency-specific properties of PAC. A mass-univariate approach was applied across all phase-amplitude frequency bins within the comodulogram at this left prefrontal location. At each bin, one-sided Wilcoxon signed-rank tests were used to assess whether normalized PAC values were significantly greater than surrogate level within each subgroup (current MDD, current control, remitted MDD, and remitted control), as well as for between-subgroup comparisons. To correct for multiple comparisons, a cluster-based approach was then implemented in which statistically significant adjacent phase-amplitude frequency bins were grouped into clusters using an 8-connected neighborhood criterion. A minimum cluster size of three contiguous bins was required for inclusion. Cluster mass was defined as the sum of Wilcoxon signed-rank test statistics within each cluster, and the maximum cluster mass was retained as the observed test statistic. Statistical significance was assessed using a nonparametric cluster-based permutation test with 5,000 permutations. For each permutation, participants were randomly reassigned to subgroups while preserving original subgroup sizes, and the full mass-univariate testing procedure was repeated across all phase-amplitude frequency bins. For each permutation, the maximum cluster mass was extracted to generate an empirical null distribution. Cluster-level p-values were computed as the proportion of permutations in which the maximum cluster mass exceeded the observed cluster mass, thereby controlling the family-wise error rate across the PAC frequency space.

## Results

There was no significant difference in sex distribution (χ²(3) = 1.22, p = 0.747) and age (F(3, 142) = 1.81, p = 0.148) across the four subgroups (Table 1). Within the current-MDD families, individuals with current MDD reported markedly higher general depression (IDAS-II) than their unaffected siblings (t(17) = 8.45, p < 0.001). Within the remitted-MDD families, individuals with remitted MDD also showed higher general depression relative to their never-depressed siblings (t(54) = 3.89, p < 0.001). In addition, negative affect (t(17) = 1.88, p = 0.076) and positive affect (t(17) = -1.43, p = 0.168) subscales of PANAS did not differ between siblings in the current-MDD families. Within the remitted-MDD families, individuals with remitted MDD showed higher negative affect relative to their never-depressed siblings (t(54) = 2.34, p = 0.022), while their differences in positive affect were not significant (t(54) = -0.27, p = 0.781).

### Cluster-based statistics of topographic PAC

#### Current-MDD families

Among individuals with current MDD, delta-beta_low_ PAC was significantly elevated in an electrode cluster over the right/central occipital and right parietal regions including channels O1, Oz, TP8, P2, P6, P8, P10, PO8, PO4, and O2 (*p* < 0.001; Figure 1A). Among the siblings of these current-MDD individuals, a significantly elevated delta-beta_low_ PAC was observed in an electrode cluster over the right/central occipital and right parietal regions at PO3, Oz, POz, P6, P8, PO8, and PO4 electrodes (*p* = 0.0012; Figure 1B). Siblings in these families did not differ on delta-beta_low_ coupling (Figure 1C).

**Figure 1.**
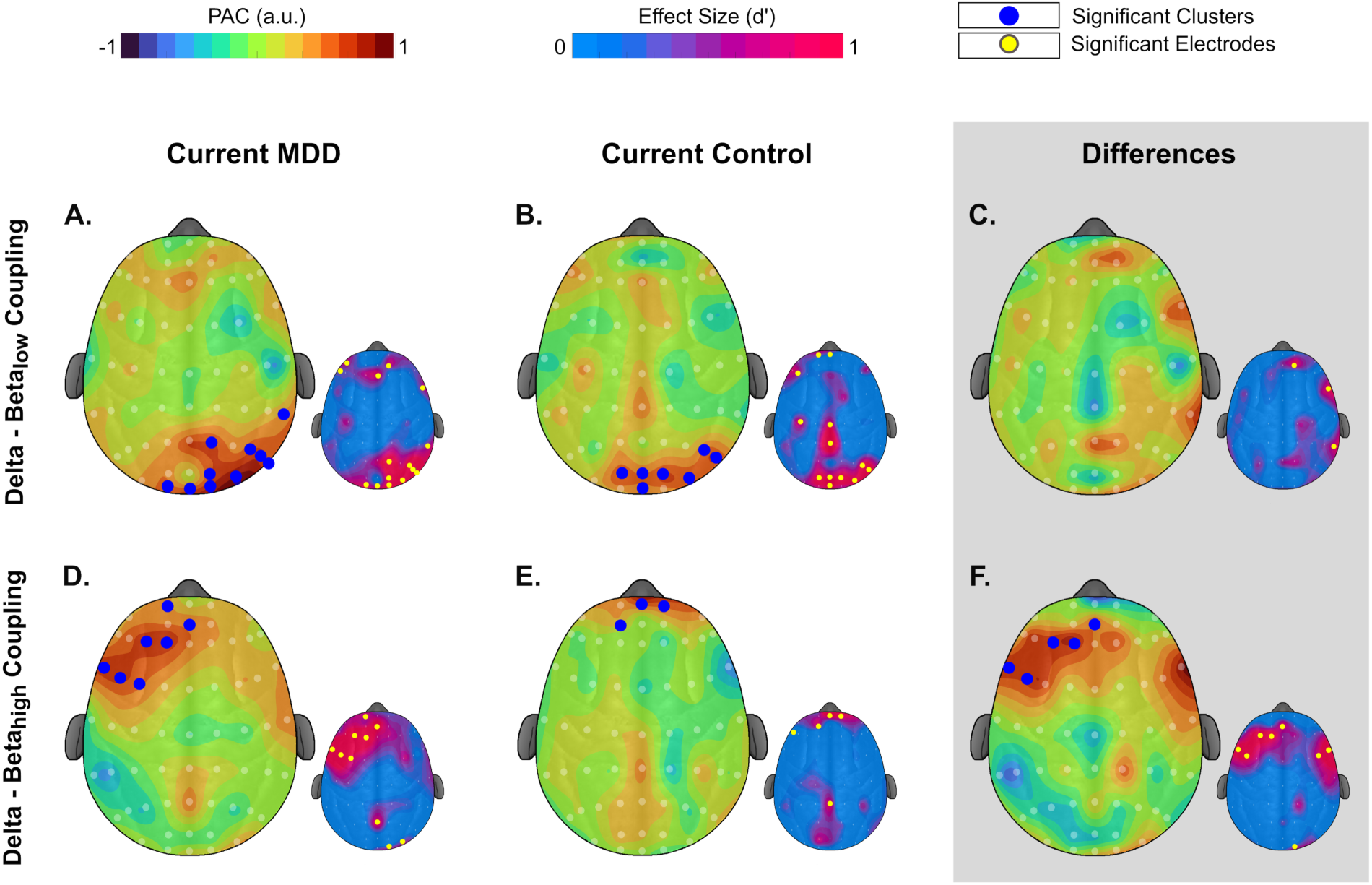
Delta-beta phase-amplitude coupling (PAC) in the current-MDD families. The larger headplots show PAC in arbitrary units (a.u.; values expressed on a relative numerical scale without standardized physical units) between delta (2-4 Hz) and beta_low_ (13-20 Hz) frequencies (top row) or beta_high_ (20-30 Hz) frequencies (bottom row). The left column depicts PAC for individuals with current MDD, the middle column depicts PAC for their non-depressed siblings, and the right column depicts the average difference within sibling pairs. Blue dots indicate electrodes with significantly elevated PAC (p < 0.05) belonging to a significant electrode cluster (p < 0.05) after cluster correction. The smaller headplots show the corresponding effect sizes in Cohen’s d units before cluster correction. Yellow dots indicate individual electrodes where PAC values are significant (p < 0.05) before cluster correction relative to the null distribution (left and middle columns) or between the sibling pairs (right column).

Delta-beta_high_ PAC was significantly elevated among individuals with current MDD over the left/central frontal cortex, with a cluster consisting of Fp1, F1, F3, FT7, FC5, FC3, and AFz electrodes (*p* < 0.001; Figure 1D). Among the siblings of these current-MDD individuals, a small cluster with elevated delta-beta_high_ PAC was observed at AF3, Fpz, and Fp2 electrodes (*p* < 0.001; Figure 1E). Individuals with current MDD had significantly greater delta-beta_high_ PAC than their siblings at the left/central prefrontal cortex (electrodes F1, F3, FC5, FC3, and AFz; *p* < 0.001; Figure 1F).

#### Remitted-MDD families

Among remitted-MDD families, delta-beta_low_ PAC was significantly elevated in the occipito-parietal regions in remitted-MDD individuals at an electrode cluster consisting of P5, PO7, PO3, O1, Oz, POz, Pz, and CP2 electrodes (p < 0.001; Figure 2A). In the siblings of remitted-MDD individuals, delta-beta_low_ PAC was also elevated in occipital and frontal regions. The significant occipital cluster comprised O1, Oz, POz, PO8, and O2 (p = 0.037), and the significant frontal cluster comprised Fp2, AFz, Fz, F2, F4, and FC2 (p = 0.048) based on the cluster-based permutation analysis (Figure 2B). Siblings in these families did not differ on delta-beta_low_ PAC (Figure 2C).

**Figure 2.**
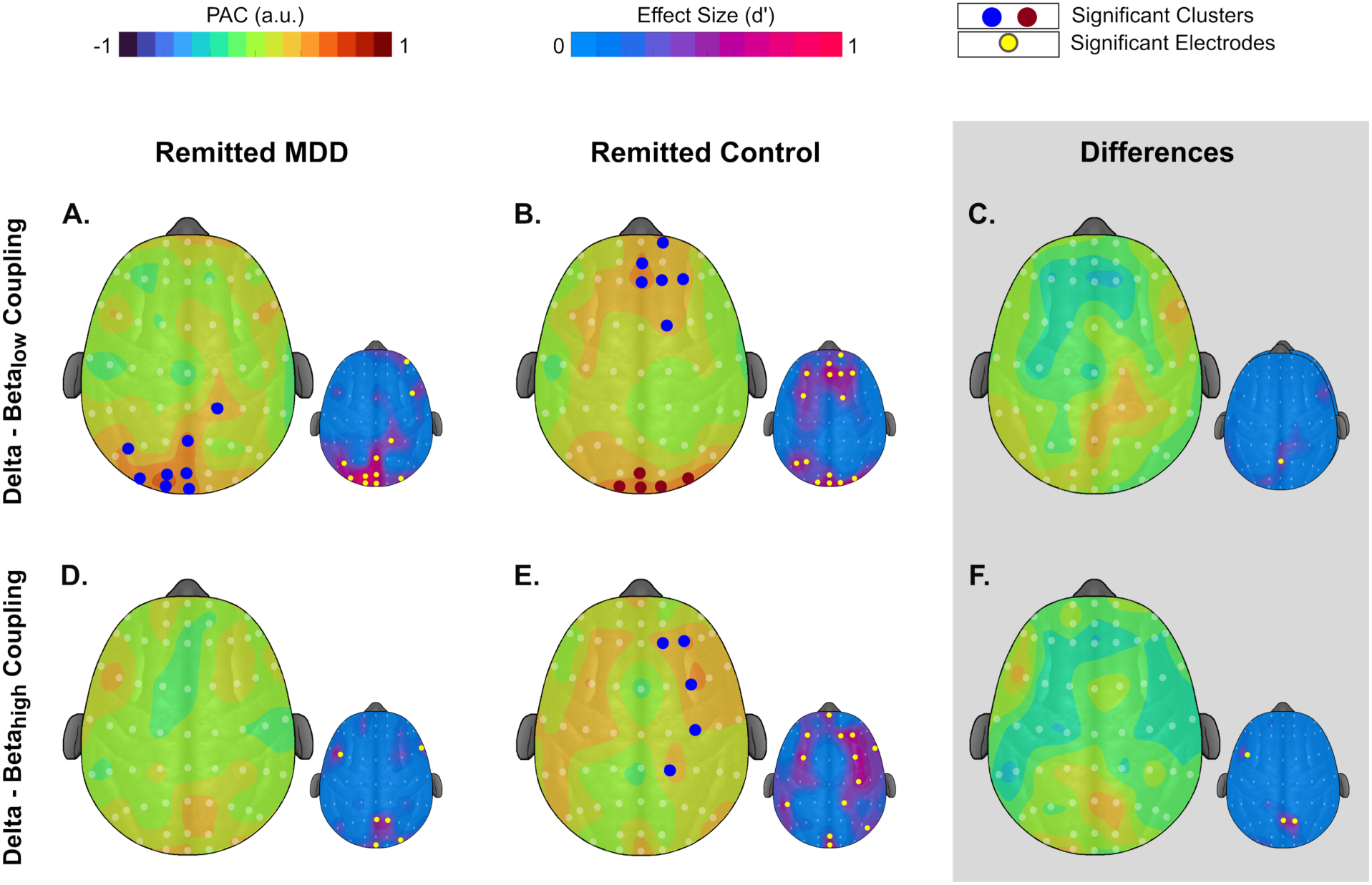
Delta-beta phase-amplitude coupling (PAC) in remitted-MDD families. The larger headplots show PAC in a.u. between delta (2-4 Hz) and beta_low_ (13-20 Hz) frequencies (top row) or beta_high_ (20-30 Hz) frequencies (bottom row). The left column depicts PAC for individuals with remitted MDD, the middle column depicts PAC for never-depressed control siblings, and the right column depicts the average differences within the sibling pairs. Blue and red dots indicate electrodes with significant PAC (p < 0.05) belonging to distinct significant clusters after cluster correction (p < 0.05). Blue dots depict the first cluster and red dots depict the second cluster. Smaller headplots show the corresponding effect sizes in Cohen’s d units before cluster correction. Yellow dots indicate individual electrodes where PAC values are significant (p < 0.05) before cluster correction relative to the null distribution (left and middle columns) or between the sibling pairs (right column).

Individuals with remitted MDD did not exhibit any significant delta-beta_high_ PAC cluster (Figure 2D). The siblings of remitted-MDD individuals exhibited significantly elevated levels of delta-beta_high_ PAC in an electrode cluster over the right frontal and right/central parietal regions, including F2, F4, FC4, C4, and CP2 electrodes (p = 0.033; Figure 2E). Siblings in these families did not differ on delta-beta_high_ PAC (Figure 2F).

As a follow-up exploratory analysis, we examined whether sibling-wise PAC differences correlated with clinical measures of depression. Results of these analyses are provided in the Supplementary Materials.

### Cluster-based statistics of PAC comodulogram

The observed left prefrontal delta-beta coupling effect was further evaluated to identify the phase and amplitude frequency ranges contributing to this pattern. The results of the first-level mass-univariate analysis revealed that, in the current-MDD families, delta-beta PAC was significant in 35 frequency bins for current-MDD individuals (Figure 3A) and in 7 frequency bins for their siblings (Figure 3B). Additionally, the difference between current-MDD individuals and their control siblings was significant in 33 frequency bins (Figure 3C). These significant bins are indicated by black contours in Figure 3. In remitted-MDD families, delta-beta PAC was not significant in any frequency bins for remitted-MDD individuals (Figure 3D), while it was significant in 9 frequency bins for their siblings (Figure 3E). No significant differences were observed between remitted-MDD individuals and their siblings (Figure 3F). Significant bins are again highlighted with black contours in Figure 3. The second-level cluster-based permutation analysis of the sibling-wise differences for the current-MDD families showed significant clusters with higher PAC in the current-MDD individuals relative to their control siblings (p = 0.0198). No significant PAC clusters were detected for remitted-MDD individuals relative to their never-depressed siblings (p = 1.000).

**Figure 3.**
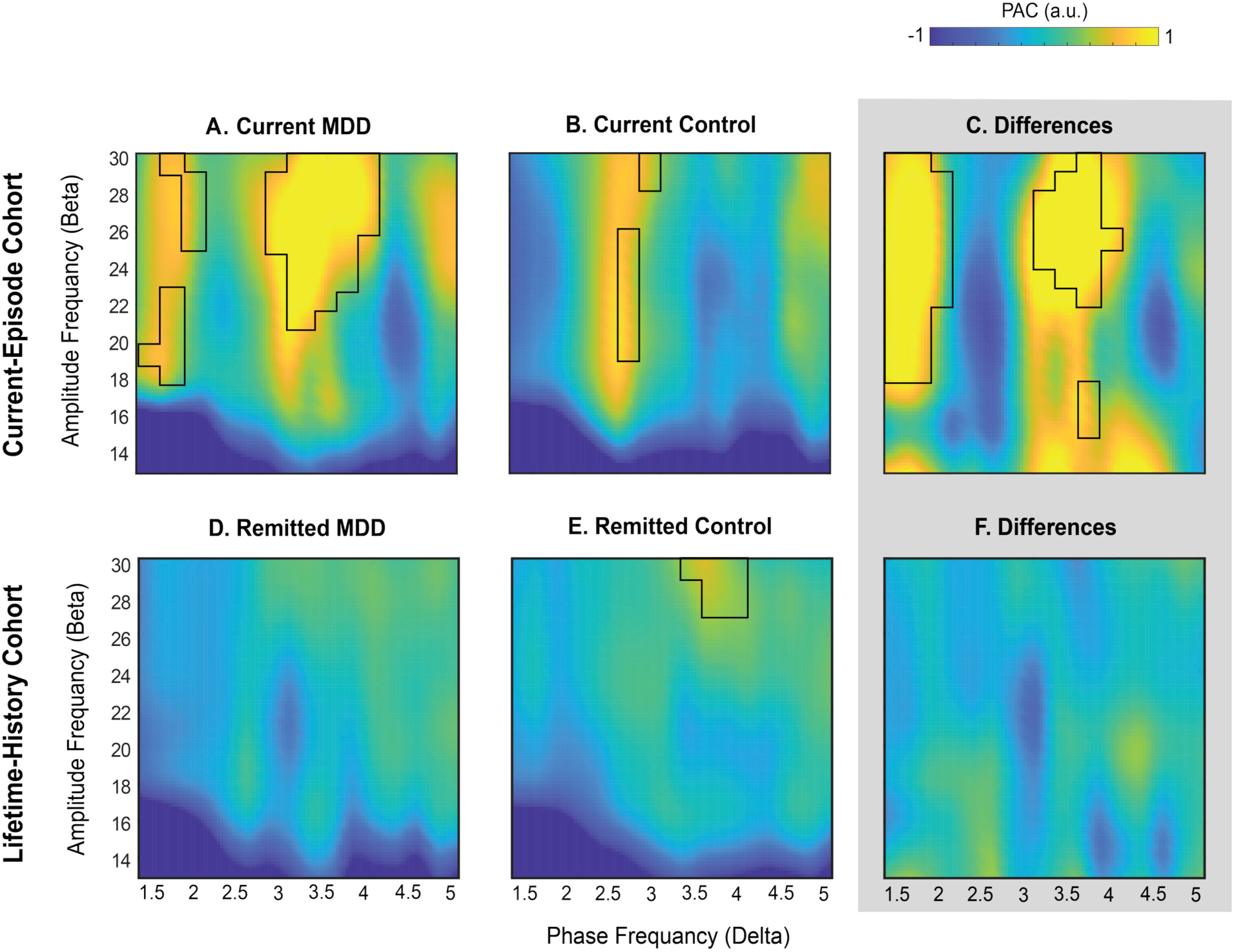
Delta-beta phase-amplitude coupling (PAC) at the left prefrontal cortex. The plots show PAC between the phase of slow oscillations (1.25-5 Hz) and the amplitude of fast oscillations (13-30 Hz) for current-MDD families (top row) and remitted-MDD families (bottom row). For each row, the left plots show PAC in siblings with current MDD (top left) and remitted MDD (bottom left), the middle plots show PAC in the siblings of current MDD (top middle) and siblings of remitted MDD (bottom middle), and the right plots show the differences between siblings in that row. Significant differences in PAC are highlighted by black contours surrounding the corresponding bins (p < 0.05). The color bar represents PAC magnitude in arbitrary units (a.u.).

### Follow-up cluster-based analysis of topographic PAC independent of family

As a follow-up analysis, we reclassified participants into three groups irrespective of their family ties: current MDD (n = 18), remitted MDD (n = 65), and never-depressed control (n = 63). We subsequently assessed delta-beta PAC across these groups to determine whether the pattern observed in the primary analyses differed between current, remitted, and never-depressed individuals.

All three groups showed elevated delta-beta_low_ PAC in the right/central occipito-parietal region. The cluster-based permutation analysis showed a significant electrode cluster consisting of O1, Oz, TP8, P2, P6, P10, PO8, PO4, and O2 electrodes in the current-MDD group (p < 0.001; Figure 4A) and a significant electrode cluster in the occipito-parietal region consisting of P5, P9, PO7, PO3, O1, Oz, POz, Pz, CPz, CP2, PO8, and PO4 in the remitted-MDD group (p < 0.001; Figure 4B). In the never-depressed control group, the significant cluster consisted of O1, Oz, POz, PO8, and O2 electrodes (p = 0.005; Figure 4C). The remitted-MDD group had higher delta-beta_low_ PAC than the never-depressed control group in one occipito-parietal cluster of PO3, Pz, CPz, and CP2 electrodes (p < 0.001; Figure 4F). The other between-group comparisons were non-significant and are presented in Supplementary Materials (Figure S4).

**Figure 4.**
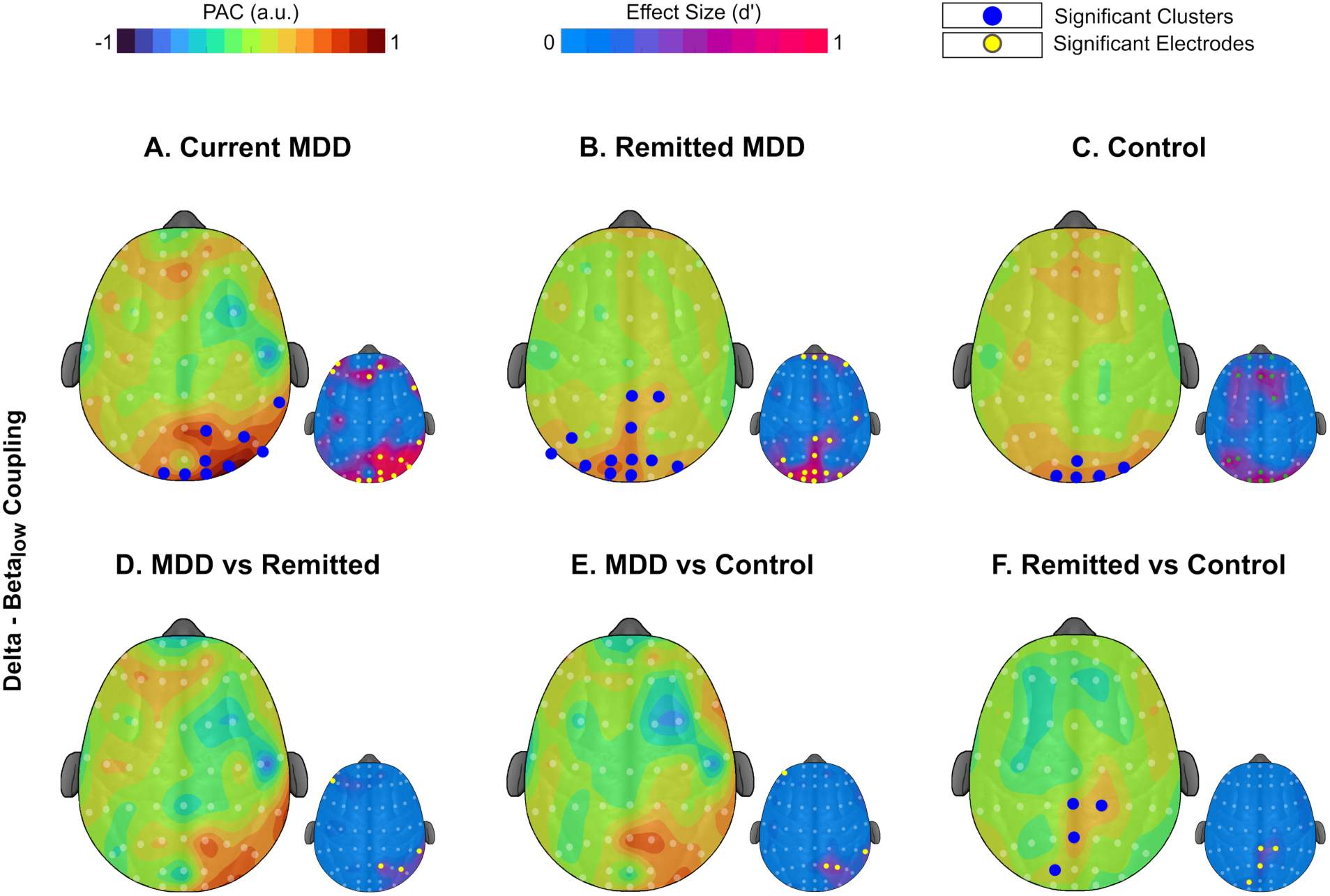
Delta-beta_low_ phase-amplitude coupling (PAC) in current MDD, remitted MDD, and never-depressed control groups (top row, in left to right columns) as well as group differences (bottom row). The larger headplots show PAC values between delta (2-4 Hz) and beta_low_ (13-20 Hz) frequencies. Blue dots indicate electrodes with significantly elevated PAC (p < 0.05) belonging to a significant electrode cluster (p < 0.05) after cluster correction. The smaller headplots show the corresponding effect sizes in Cohen’s d units before cluster correction. Yellow dots indicate individual electrodes where PAC values are significant (p < 0.05) before cluster correction relative to the null distribution (left and middle columns) or between the sibling pairs (right column).

Delta-beta_high_ coupling was significantly elevated in the current-MDD individuals in the left/central frontal region at an electrode cluster consisting of Fp1, F1, F3, FT7, FC5, FC3, and AFz electrodes (p < 0.001; Figure 5A). There was no significant electrode cluster for the remitted-MDD and never-depressed control groups. Current-MDD individuals exhibited significantly higher delta-beta_high_ PAC than remitted-MDD individuals within a cluster in the left/central region comprising Fp1, F1, F3, FT7, FC5, AFz, and Fz electrodes (p < 0.001; Figure 5D). Current-MDD individuals also exhibited significantly higher delta-beta_high_ PAC than never-depressed controls in a similarly located cluster (F1, F3, FT7, and FC5; p < 0.001; Figure 5E). The other between-group comparisons are presented in Supplementary Materials (Figure S4).

**Figure 5.**
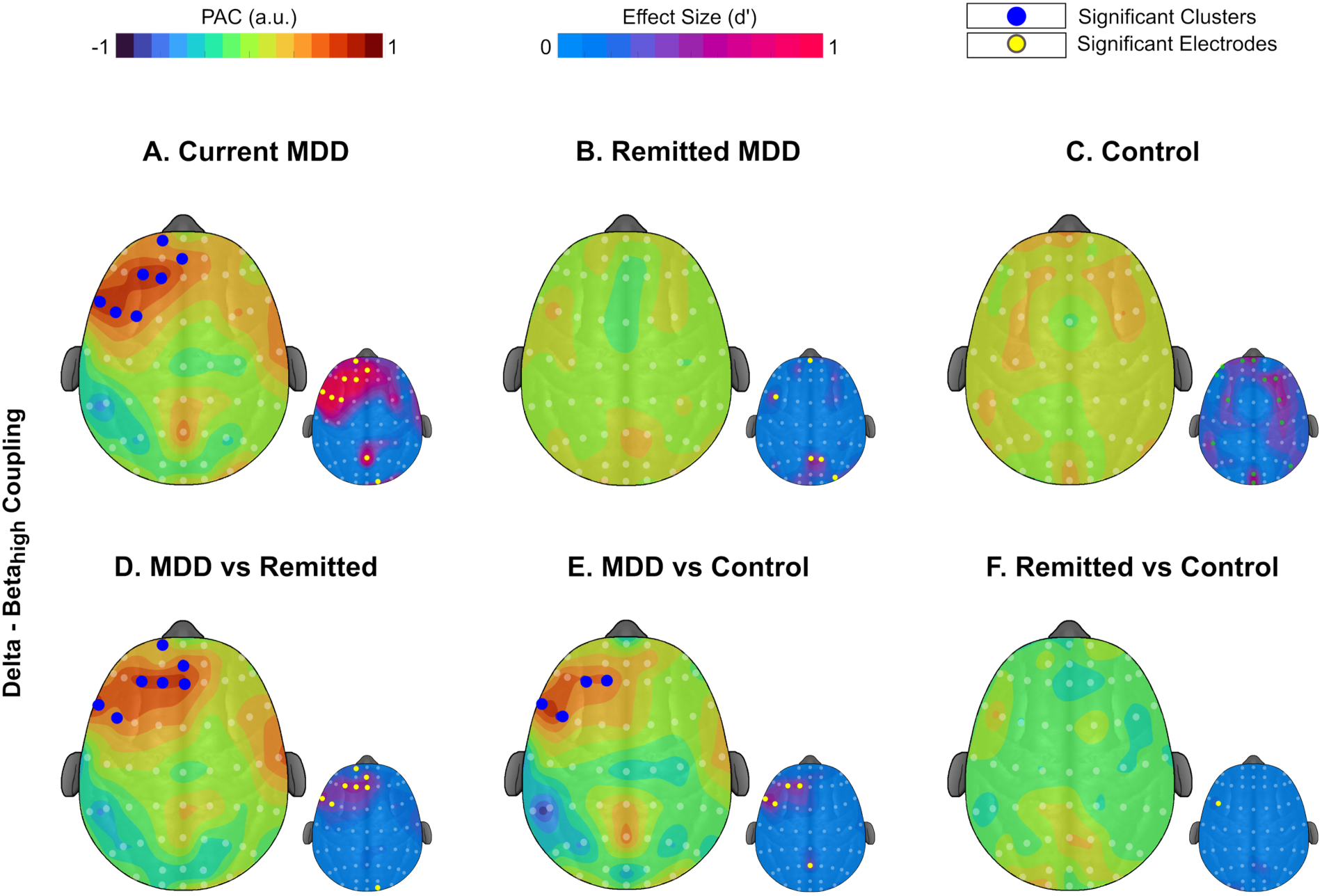
Delta-beta_high_ phase-amplitude coupling (PAC) in current MDD, remitted MDD, and never-depressed control groups (upper row, in left to right columns) as well as group differences (lower row). The larger headplots show PAC values between delta (2-4 Hz) and beta_high_ (20-30 Hz) frequencies. Blue dots indicate electrodes with significantly elevated PAC (p < 0.05) belonging to a significant electrode cluster (p < 0.05) after cluster correction. The smaller headplots show the corresponding effect sizes in Cohen’s d units before cluster correction. Yellow dots indicate individual electrodes where PAC values are significant (p < 0.05) before cluster correction relative to the null distribution (left and middle columns) or between the sibling pairs (right column).

## Discussion

Using a familial sibling-pair design, the present study investigated the spatial and spectral characteristics of delta-beta phase-amplitude coupling (PAC) in depression, with a particular focus on whether PAC alterations reflect state-versus trait-related features of depression. In the current-MDD families, resting-state delta-beta_high_ PAC was elevated over the left and central frontal cortex in those currently in a depressive episode relative to their asymptomatic siblings. In the remitted-MDD families (i.e., both siblings were currently asymptomatic, but one had a past depressive episode), no such between-sibling difference emerged. A reanalysis that regrouped participants irrespective of sibling pairing reproduced the left and central prefrontal elevation specifically in the current-MDD group relative to both remitted and control participants. Together, these findings indicate that elevated left-central frontal delta-beta PAC tracks the state of active depression rather than a stable or familial trait predisposing to the disorder.

Changes in delta oscillation amplitudes are thought to reflect activity in subcortical networks involved in affective and motivational processes (Knyazev, 2007; Nelson et al., 2018), whereas changes in beta oscillations are thought to reflect activity in local cortical regions associated with attentional control and regulation of negative affect (Engel & Fries, 2010; Güntekin & Başar, 2010; Lopes da Silva, 1991). Accordingly, delta-beta coupling may serve as a proxy of functional communication between local cortical control regions and subcortical limbic structures. This interaction is thought to index top-down cognitive processes involved in monitoring and inhibiting emotional arousal and reactivity (Brooker et al., 2021; Knyazev, 2007, 2007; Miskovic et al., 2011; Putman, 2011). Similarly, stronger delta-beta coupling has been suggested to reflect rigid and inflexible neural patterns, which may manifest in depression as ruminative and hypervigilant cognitive patterns (P. Liu et al., 2018). For example, elevated levels of frontal delta-beta PAC during stress anticipation were associated with enhanced negative perceptual biases and stress reactivity (Montijn et al., 2023). Taken together, these findings support the view that delta-beta coupling reflects the current state of emotional disorders and may hold promise as a biomarker to be used in assessment and treatment of MDD.

The left-prefrontal topography effect aligns with a long line of evidence implicating the prefrontal cortex (PFC; particularly left PFC) in depression (Shankman & Klein, 2003). This region supports cognitive control, executive function, and emotion regulation (H. Liu et al., 2016; Pan et al., 2019; Pizzagalli & Roberts, 2022; Zhang et al., 2022). Several studies have reported increased activity asymmetry in the right and the left PFC in depression during rest (Coan & Allen, 2004; Plewnia et al., 2015; Spironelli et al., 2020) and emotional processing (Shankman et al., 2007, 2013), mostly reflected in increased left-frontal EEG alpha power (Chen et al., 2026; Davidson, 1998; Stewart et al., 2014). PFC activity is also a primary target for therapeutic interventions such as transcranial magnetic stimulation (Trapp et al., 2025). However, our results suggest that the left prefrontal cortex in depression at rest is not *inhibited* but rather *engaged* in a coupling regime that fails to translate into adaptive affective regulation. Supporting our findings, a multi-case study in bipolar disorder observed reduced left-frontal delta-beta coupling after three weeks of lamotrigine treatment, and a positive association between coupling strength and depression severity (Kesebir et al., 2019). Others have found greater delta-beta coupling in adolescents with more severe clinician-rated depression undergoing CBT, although self-reported severity showed an effect in the opposite direction (Venanzi et al., 2024). The present study aims to clarify these prior findings using a well-powered, within-family discordant design that reduces the effects of genetic and environmental confounds shared between siblings. Notably, the direction of our resting-state PAC effect runs counter to what is observed for delta-beta PAC with anhedonia during reward-based decision-making (Riddle et al., 2022). This dissociation likely signals that the delta-beta PAC indexes different processes at rest versus during goal-directed tasks. Thus, elevated frontal delta-beta coupling in current MDD may reflect inflexible or maladaptive prefrontal engagement with subcortical structures, rather than suppressed frontal activity.

Elevated delta-beta PAC was specific to current MDD and was absent following remission, with remitted individuals showing levels comparable to never-depressed individuals. In addition to being familial, a vulnerability trait marker is proposed to be present before, during, and after an episode of psychopathology (Zubin & Spring, 1977). Thus, the absence of elevated delta-beta PAC in remitted individuals suggests that this alteration is more consistent with a state-related marker of active depression than a persistent trait vulnerability. A substantial proportion of individuals who experience a depressive episode will experience subsequent episodes, with recurrent depression presenting an ongoing challenge for decisions regarding discontinuation or continuation of maintenance treatment. Accordingly, objective and reliable biomarkers that can distinguish active depression from remission could have clinical value, particularly in informing the assessment of residual vulnerability and relapse risk. Our findings suggest that delta-beta PAC may have potential as a neurophysiological marker for differentiating an active MDD episode from the remitted state. However, longitudinal studies are needed to determine whether delta-beta PAC changes systematically with symptom onset and remission and whether these changes have utility for monitoring treatment response or predicting relapse. We emphasize, however, that “state” here means the presence of an active episode rather than within-episode clinical severity. With that caveat, the result identifies a neurophysiological state biomarker.

In our study, altered coupling was specific to delta-beta_high_ PAC, but not delta-beta_low_ PAC, in current-MDD participants. One study examining the effects of sleep deprivation on state anxiety reported a significant positive correlation between delta-beta_high_ PAC and anxiety severity in frontal and occipital regions, whereas no such association was observed for delta-beta_low_ PAC (Hao et al., 2025). Several studies have reported that those with MDD show greater elevations in the beta_high_ range than in the beta_low_ range compared to healthy controls (Johnson et al., 2026; X. Liu et al., 2024). In addition, beta_high_ activity plays a stronger role in discriminating between emotional states than beta_low_ activity (De Filippi et al., 2021b). We identified elevated delta-beta_low_ PAC over occipito-parietal cortex in every subgroup we examined, but with no significant between-subgroup differences. We interpret this posterior coupling as a non-specific feature of the eyes-closed resting state rather than a feature of psychopathology.

The study has several strengths. First, examining families of current- and remitted-MDD individuals can point to “state” vs. “trait” effects (although studies of remitted individuals may also point to scar effects (Letkiewicz et al., 2022)). Second, the sibling-discordant comparisons partially control for shared familial effects. Third, multi-level permutation statistics limit Type I error and protect against spurious findings. Limitations of the current study should also be considered. First, the study design is cross-sectional, so individuals were not followed from an active episode into remission. Second, about 14% of participants were taking psychotropic medication, which we did not model due to sample size, and which likely introduced variance. Third, the study did not include individual MR imaging, which prevents reliable EEG source localization. These constraints point to several next steps. One key next step is to use frequency-specific neuromodulation. This includes transcranial alternating current stimulation phase-locked to delta over the left prefrontal cortex and TMS protocols engaging delta-beta coupling. Showing that such protocols result in transitions from current to remitted states could offer strong evidence of delta-beta PAC as a causal biomarker of MDD. Furthermore, although prior work in anxiety has linked delta-beta correlation to a network involving the orbitofrontal and anterior cingulate cortices (Knyazev, 2011), future simultaneous EEG-fMRI studies should investigate the specific resting-state networks underlying delta-beta PAC in depression. Finally, comparison to other internalizing disorders, particularly anxiety, would clarify whether the prefrontal state signature is specific to MDD or shared across emotion-regulation pathologies.

Taken together, eyes-closed resting-state delta-beta_high_ phase-amplitude coupling over the frontal cortex is pathologically elevated in individuals with current MDD but not in their currently unaffected siblings or in individuals with remitted MDD. This biomarker tracks with state depressive symptoms and is independent of familial trait factors. Thus, delta-beta_high_ PAC represents a physiologically defined target for future treatment studies and rhythmic neuromodulation therapies.

## Data Availability

All data produced in the present study are available upon reasonable request to the authors

## Acknowledgments

Research reported in this publication was supported by the National Institutes of Health under award numbers R01MH134369 and R01MH098093 (awarded to Stewart Shankman), T32NS047984 (awarded to Mina Elhamiasl), and K23MH136421 (awarded to Brent Rappaport). The content is solely the responsibility of the authors and does not necessarily represent the official views of the National Institutes of Health.

