## Supplementary Materials for "Prefrontal EEG Delta-Beta Coupling Marks Depressed State in Adult Siblings"

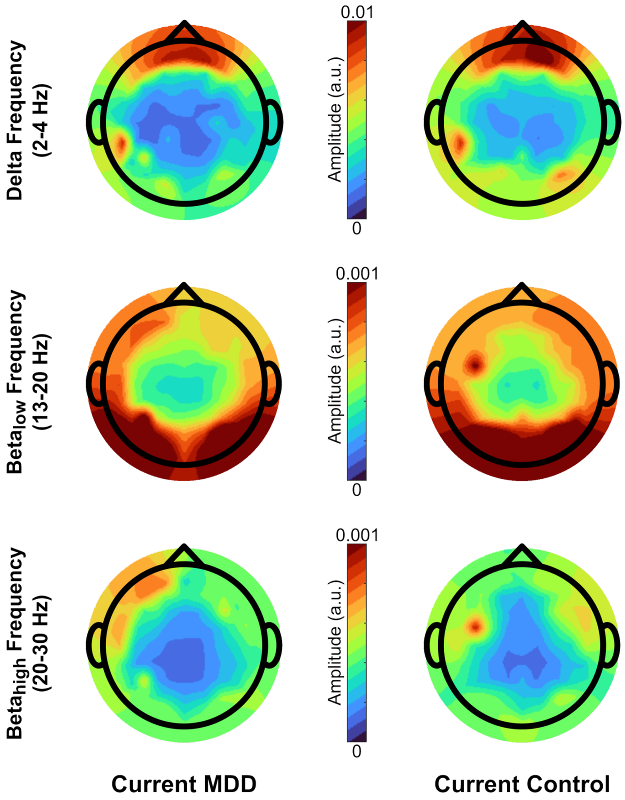


**Figure S1.** Distribution of normalized power for phase and amplitude frequencies in the current-MDD families. The headplots show normalized power for delta (2-4 Hz), beta$_{low}$ (13-20 Hz), and beta$_{high}$ (20-30 Hz) frequencies over all electrodes. The left column indicates the distribution in siblings with current MDD, and the right column indicates the distribution in their currently non-depressed siblings. The color bar represents power magnitude.

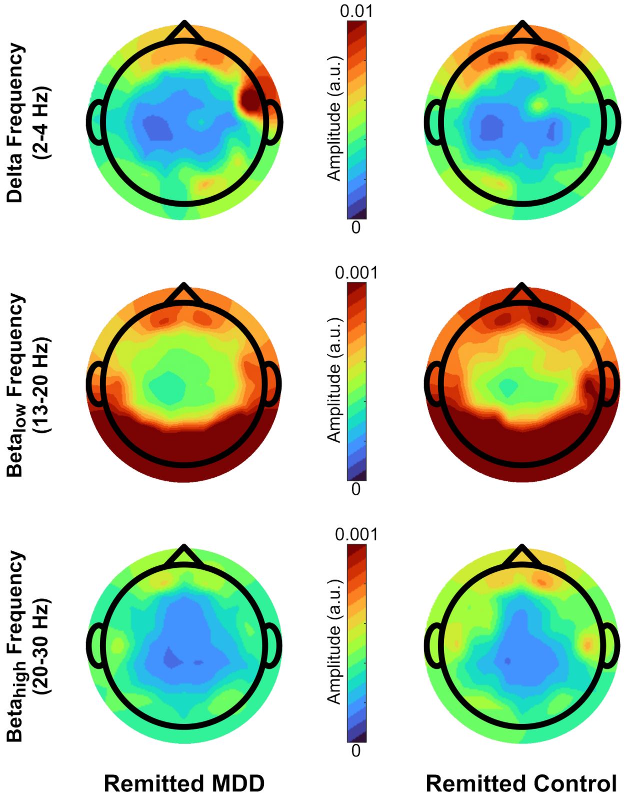


**Figure S2.** Distribution of normalized power for phase and amplitude frequencies in the remitted-MDD families. The headplots show normalized power for delta (2-4 Hz), $beta_{low}$ (13-20 Hz), and $beta_{high}$ (20-30 Hz) frequencies over all electrodes. The left column indicates the distribution in siblings with remitted MDD, and the right column indicates the distribution in their never-depressed siblings. The color bar represents power magnitude.

*Associations Between Delta-Beta PAC and Clinical Measures*

Given the significant subgroup differences in frontal delta-beta PAC in the current-MDD families, we performed an exploratory post hoc analysis of the unidirectional relationships between the mean delta-beta PAC values within the significant frontal electrode cluster and three key clinical measures: the general depression subscale of the IDAS-II (Watson et al., 2012) and the positive and negative affect subscales of the PANAS (Watson et al., 1988).

*Regression analysis:* Within-sibling-pair differences were computed for each variable (*n* = 73 pairs). Three missing IDAS-II values and two missing PANAS values were imputed with the corresponding subgroup mean. At the first level, we fitted a generalized linear mixed-effects (GLME) model with the delta-beta PAC difference as the dependent variable and the three clinical-score differences, together with the positive-by-negative affect interaction, as fixed-effect predictors. Age difference (in years), sex pairing (coded as 1 - two males, 2 - two females, 3 - a male in the MDD subgroup with a female in the control subgroup, and 4 - the inverse pairing), and their interaction were entered as random effects. Participants from both current-MDD and remitted-MDD families were pooled for this exploratory analysis to maximize statistical power and evaluate symptom-PAC relationships across the full sample. At the second level, for the clinical measures that reached significance in the GLME, we computed Spearman rank-order correlations, first on all available data and then on the cleaned data. Cleaning consisted of removing the imputed data points together with any value lying more than 3 SD from the variable's mean, to strictly satisfy the assumptions of the correlation analysis.

*Regression Results:* The GLME model revealed a significant regression of negative affect on PAC differences ($F_{1,65} = 4.96$, p = 0.029), whereas positive affect ($F_{1,65} = 2.28$, p = 0.14), the positive-by-negative interaction ($F_{1,65} = 1.88$, p = 0.17), and depressive symptoms ($F_{1,65} = 0.52$, p = 0.47) were not significant. Follow-up Spearman correlations confirmed a significant unidirectional association between negative affect and delta-beta PAC (r = 0.24, p = 0.04), but not between PAC and either positive affect or depressive symptoms (both p > 0.2). Even when removing possible outliers, negative affect remained significantly correlated with PAC (r = 0.25, p = 0.038). Results are presented in Figure S3.

Within-sibling differences in left prefrontal delta-beta PAC showed a weak, significant association with negative affect and no association with positive affect or depressive symptoms. The difference between the robust categorical group effect and weak dimensional associations is consistent with several non-exclusive interpretations, including restricted symptom range within the subgroups, the difference in timescale between resting EEG measurement and psychometric self-report, and confounding factors like medication. This dimensional analysis can inform future, appropriately powered studies examining whether delta-beta PAC tracks MDD symptom severity.

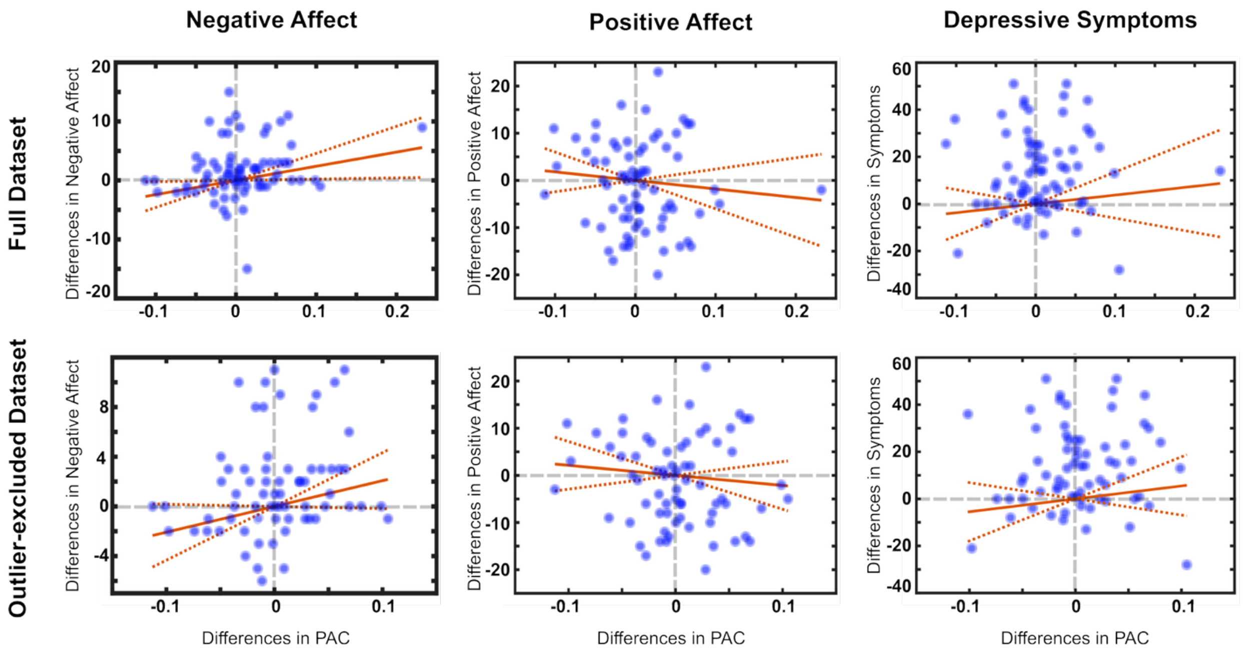


**Figure S3.** Scatterplots of within-sibling-pair differences in left central-frontal delta-beta phase-amplitude coupling (PAC), plotted against within-pair differences in negative affect (left column), positive affect (middle column), and depressive symptoms (right column). The upper row shows all available data points ($n$ = 73 sibling pairs). The lower row redraws the same data after removing the imputed values and putative outliers. Plots showing a significant unidirectional relationship ($p < 0.05$ in both the GLME and the Spearman correlation) are highlighted with a thicker border. The gray dashed line indicates the zero line.

*Cluster-Based Permutation of PAC Across Electrodes in Three Groups: Additional Group Comparisons*

The additional between-group cluster-based permutation analysis of delta-beta$_{low}$ as well as delta-beta$_{high}$ PAC across all electrodes showed that differences were significant only between never-depressed control vs. current-MDD groups. For delta-beta$_{low}$, PAC was higher in control than current-MDD participants at one cluster of FC4, FC2, and C6 ($p < 0.001$; Figure S4B). For delta-beta$_{high}$, PAC was higher in control than current-MDD individuals at one cluster of T7, CP5, and P3 ($p < 0.001$; Figure S4E).

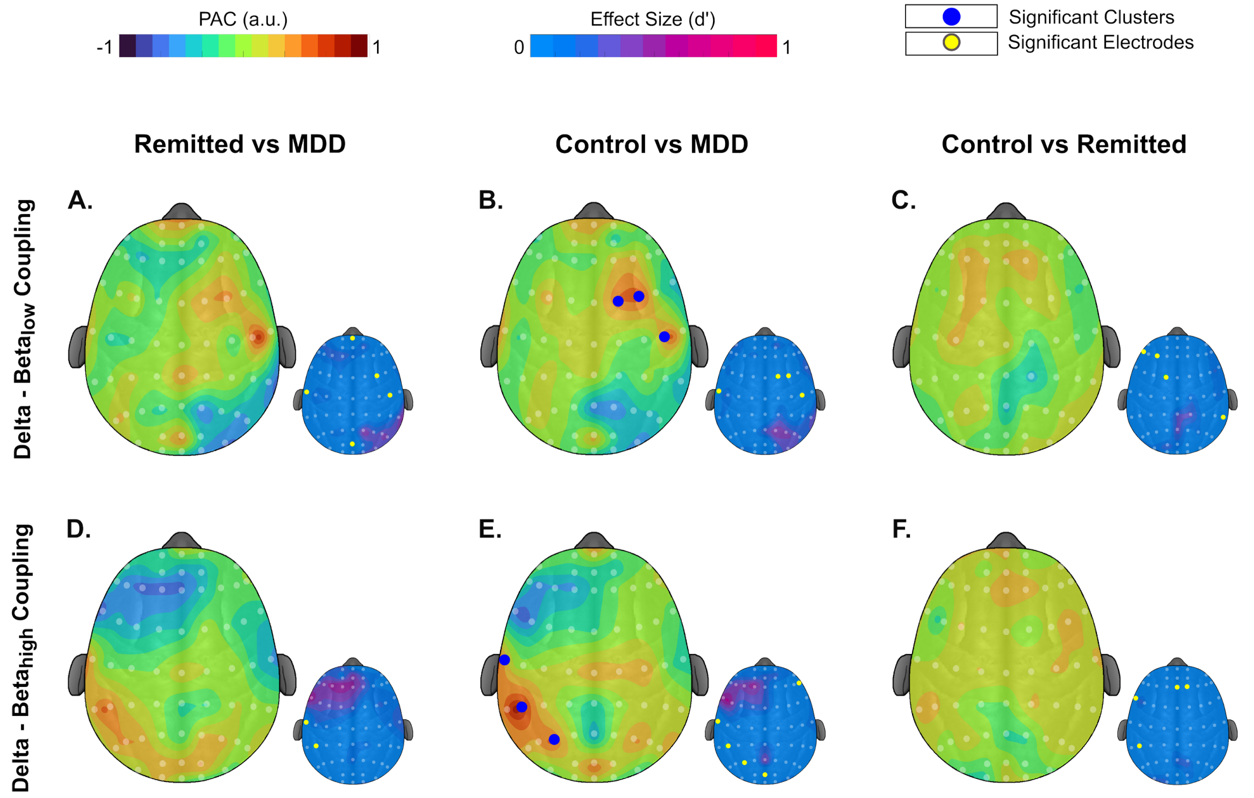


**Figure S4.** Delta-beta phase-amplitude coupling (PAC) differences between current MDD, remitted MDD, and never-depressed groups. The larger headplots show PAC between delta (2-4 Hz) and beta$_{low}$ (13-20 Hz) frequencies (top row) as well as delta (2-4 Hz) and beta$_{high}$ (20-30 Hz) frequencies (bottom row). The left column reports remitted MDD vs. current MDD PAC differences, the middle column reports never-depressed vs. current-MDD individuals PAC differences, and the right column shows never-depressed individuals vs. remitted MDD PAC differences. Blue dots indicate electrodes with significant PAC ($p < 0.05$) belonging to significant clusters ($p < 0.05$) after cluster correction. Yellow dots indicate individual electrodes where PAC values are significant ($p < 0.05$) before cluster correction relative to the null distribution (left and middle columns) or between the sibling pairs (right column).
